# BNT162b2 vaccine booster dose protection: A nationwide study from Israel

**DOI:** 10.1101/2021.08.27.21262679

**Authors:** Yinon M. Bar-On, Yair Goldberg, Micha Mandel, Omri Bodenheimer, Laurence Freedman, Nir Kalkstein, Barak Mizrahi, Sharon Alroy-Preis, Nachman Ash, Ron Milo, Amit Huppert

**Affiliations:** Weizmann Institute of Science, Israel; Technion - Israel Institute of Technology, Israel; The Hebrew University of Jerusalem, Israel; Israel Ministry of Health, Israel; The Gertner Institute for Epidemiology & Health Policy Research, Sheba Medical Center, Israel; KI Research Institute, Kfar Malal, Israel; Tel Aviv University, Israel

## Abstract

**Background:** On July 30, 2021, a third (booster) dose of the Pfizer BNT162b2 vaccine was approved in Israel for individuals 60 years or older who had been fully vaccinated (i.e., received two doses) at least five months previously. Here, we estimate the reduction in relative risk for confirmed infection and severe COVID-19 provided by the booster dose.

**Methods:** 1,144,690 individuals aged 60y and older who were eligible for a booster dose were followed between July 30 and August 22, 2021. We defined dynamic cohorts where individuals initially belong to the ‘non-booster’ cohort, leave it when receiving the booster dose and join the ‘booster’ cohort 12 days later. Rates of infection and severe COVID-19 outcomes per person-days at risk were compared between the cohorts using Poisson regression, adjusting for possible confounding factors.

**Results:** Twelve days or more after the booster dose we found an 11.4-fold (95% CI: [10.0, 12.9]) decrease in the relative risk of confirmed infection, and a >10-fold decrease in the relative risk of severe illness. Under a conservative sensitivity analysis, we find ≈5-fold protection against confirmed infection.

**Conclusions:** In conjunction with safety reports, this study demonstrates the effectiveness of a third vaccine dose in both reducing transmission and severe disease and indicates the great potential of curtailing the Delta variant resurgence by administering booster shots.

## Introduction

The rapid development of effective vaccines against SARS-CoV-2 and their deployment to the general population has been proven to be a highly successful strategy for reducing transmission and disease burden. In Israel, a swift vaccination campaign led to more than half of the population being fully vaccinated by the end of March 2021 ^1^. Consequently, COVID-19 incidence dropped from ≈900 cases per million per day in mid-January 2021 to less than 2 cases per million per day by June 2021 ^1^. Nevertheless, the emergence of new variants of concern (VOC), and specifically the Delta variant, has led to a recent infection resurgence in Israel both in infection and severe disease ^2^. There are several possible causes for the high levels of transmission of the Delta variant, including increased infectiousness of the Delta variant ^3^, waning vaccine-elicited immunity ^2,4^, and heightened immune evasion by the variant ^5^, the latter two of which directly contribute to a decrease in vaccine efficacy. Analysis of the Israeli data on the Delta outbreak indicated strong waning immunity. In an effort to address the challenge presented by the Delta variant and reduce the load on the healthcare system, Israeli authorities approved the administration of a booster dose, first to high-risk populations, on July 12, 2021, and then to the entire 60+ population, on July 30, 2021.

Initial studies have suggested that a BNT162b2 booster dose, i.e., an additional dose given to individuals who have previously received two BNT162b2 vaccine doses, increases antibody neutralization levels ∼10-fold, on average, compared to levels achieved after the second dose^6^. It is thought that an increased neutralization titer could lead to increased protection against infection and severe illness ^7^. However, in terms of real-world efficacy, the size of such an effect remains unclear. Here, we use initial data from the Israeli Ministry of Health (MOH) database on the incidence of confirmed infection and severe illness among two cohorts of individuals above 60 years of age: those who received only two vaccine doses, and those who received an additional booster dose. We use the data to quantify the protective effect that the booster dose provides against confirmed infection and severe illness.

The protection gained by the booster shot is not expected to reach its maximal capacity immediately on vaccination, but to build up over the week following vaccination ^8,9^. At the same time, during the first days after vaccination, significant behavioral changes in the booster-vaccinated population are expected (Figure S1 in the Supplementary Appendix). One such expected change is added avoidance of exposure to excess risk until the booster dose becomes effective. Another expected change is a reduced rate of testing for COVID-19 around the time of receiving the booster, as demonstrated in Figure S2 (Supplementary Appendix). Moreover, we analyzed confirmed COVID-19 infections based on the date of the positive PCR test, and testing occurs only several days following exposure. For all these reasons, it is preferable to assess the effect of the booster only after a sufficient period has passed since its administration.

## Methods

Our analysis is based on medical data from the MOH database extracted on August 24, 2021. There were 1,186,780 Israeli residents aged 60 and older who had been fully vaccinated at least five months (became fully vaccinated before March 1, 2021), and were still alive on July 30, 2021. We removed from these data individuals who: had missing gender; were abroad in August 2021; had been infected with COVID-19 before July 30, 2021; received a booster dose before July 30, 2021; or became fully vaccinated before January 16. A total of 1,144,690 individuals met the inclusion criteria for the analysis (see Figure 1). The data included vaccination dates (first, second and third doses), RT-qPCR tests (dates and results), COVID-19 hospitalization date (if relevant), demographic variables such as age, gender, and demographic group (General Jewish, Arab, ultra-Orthodox Jewish) ^10^, and clinical status (mild, severe). Severe disease was defined as: resting respiratory rate >30 breaths per minute, or oxygen saturation on room air <94%, or ratio of PaO2 to FiO2 <300 ^11^.

**Figure 1.**
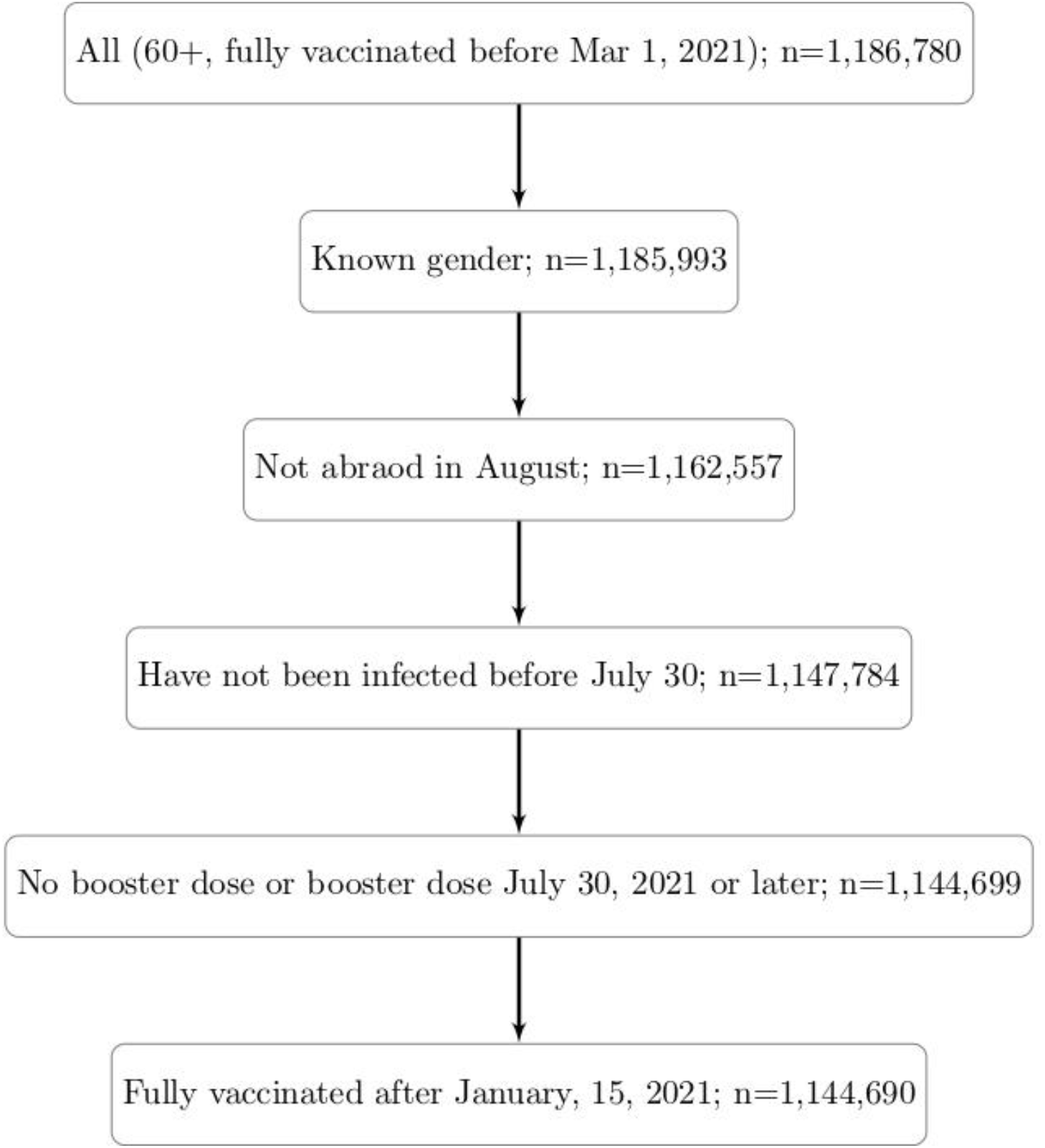
Study population. The population includes people who were fully vaccinated prior to March 1, 2021, were not abroad during August 2021, and had no documented SARS-CoV-2 PCR-positive result before July 30, 2021.

We considered 12 days as the time it took the booster dose to affect the observed number of confirmed infections. Our study period started at the beginning of the booster vaccination campaign on July 30, 2021. The end date was chosen as August 22, 2021, to minimize the effects of missing outcome data due to delays in the reporting of test results. Choosing 12 days following booster vaccination as the cutoff is scientifically justified from an immunological perspective, as studies have shown that following the booster dose, neutralization levels increase only after several days ^6^. Using confirmed infections (i.e., PCR positivity) as an outcome, there is a delay between infection and testing. For symptomatic cases, infections occur on average 5-6 days prior to testing, similar to the incubation period of COVID-19 ^12,13^.

To estimate the level of protection provided by the booster dose, we analyzed data on the incidence of confirmed infections and severe illness of two distinct cohorts: people who received two vaccine doses and the booster dose (‘booster’ cohort), and people who received only two vaccine doses (‘no-booster’ cohort). These cohorts were dynamic; individuals initially belonged to the ‘non-booster’ cohort, left it when receiving the booster dose, and joined the ‘booster’ cohort 12 days later (Figure S3 in the Supplementary Appendix) provided they did not have a confirmed infection in the interim period. We considered data on two outcomes of interest, confirmed infection and severe COVID-19, and counted the number of events of each type during the study period.

For each cohort, we calculated the incidence rate of both confirmed infection and severe COVID-19 per person-days at risk. For each person in the ‘booster’ cohort, days at risk started when entering the cohort (12 days after receiving the third dose), and ended either with the occurrence of an outcome or at the end of the study period. For the ‘no-booster’ cohort, days at risk started at the beginning of the study period (August 10, 2021), and ended either with the occurrence of an outcome, the end of the study period, or when receiving a booster dose. Since cohort membership was dynamic, many individuals contributed person-days at risk to both cohorts.

We fitted a Poisson regression (using the glm function in the R Statistical Software^)14^ to estimate the incidence rate of a specific outcome, controlling for several important covariates: age (60-69, 70-79, 80+), gender, demographic group (General Jewish, Arab, ultra-Orthodox Jewish) ^10^, and date of second vaccine dose (in half-month intervals). Since the overall incidence rate of both confirmed infection and severe COVID-19 increased exponentially during the study period, days at the beginning of the study period had lower exposure risk than days at the end. To account for growing exposure risk, we included calendar date as an additional covariate. Accounting for these covariates, we used the study cohort (‘booster’ or ‘no-booster’) as a factor in the regression and estimated its impact on the incidence rate. The effect of the booster dose is estimated as one divided by the exponent of the regression coefficient associated with the treatment cohort, which is akin to a relative risk. For reporting uncertainty around our estimate, we used the exponent of the 95% confidence limits for the regression coefficient.

As a sensitivity analysis, we compared infection rates before and after the booster dose became effective. Specifically, we repeated the Poisson regression analysis described above but compared infection rates on days 4-6 to 12+ after the booster dose. Our hypothesis was that the booster dose was not yet effective during the former period ^8^. This analysis compares different periods following booster vaccination based only on those who received the booster dose, and may reduce selection bias. On the other hand, people might perform less PCR testing and behave more cautiously with regard to virus exposure just after getting the booster vaccination (Figure S2), so we conjecture that the protection effect is underestimated in this analysis, providing a lower bound to the real effect.

To further examine the protection as a function of time from the booster dose, we fitted a Poisson regression comparing the ‘booster’ and ‘no-booster’ cohorts as above, while including each day, from day 1 up to day 24 after the booster vaccination, as a separate factor in the model. The period before receiving the booster dose (‘no-booster’ cohort) was used as the reference category. The analysis is similar to the Poisson modeling described above, with one divided by the exponents of the regression coefficients representing the protection on different days post the booster vaccination. The follow-up time for this analysis started on July 30, 2021, and ended on August 22, 2021.

As further sensitivity analyses, we applied two methods based on case-control matching. The first method was similar to that used by Dagan et. al^15^. Each person who received the booster was matched, using the same covariates as used in the Poisson regression model, to a person who had not yet received it on that date. We compared the probabilities of COVID-19 infection 12 days or more after the matching time of those receiving the booster dose and those who did not. In the second method, we matched person-days rather than individuals, ensuring that person-days in the two cohorts are comparable in terms of covariates and exposure risk. On each day we identified the group of individuals for whom 12 days or more had passed since receiving the booster dose and who had not been infected in the interim (‘booster’ cohort). We randomly matched a day of a ‘no-booster’ individual from those who had received only two vaccine doses by that same date, had not been previously infected, and had the same characteristics (age, gender, second vaccination period, and demographic group). We then calculated the risk ratio of infection and severe COVID-19 between the two groups. A detailed description of both approaches is given in Supplementary Methods 1 in the Supplementary Appendix.

## Results

The baseline characteristics of both cohorts are shown in Table 1. As our primary analysis adjusts for person-days at risk, and since individuals contribute days to both cohorts, we compare characteristics according to person-days at risk. There are about 4.0 million person-days in the ‘no-booster’ cohort with 3,473 confirmed infections and 330 cases of severe COVID-19, and about 3.4 million person-days in the ‘booster’ cohort with 313 confirmed infections and 32 cases of severe COVID-19. The ‘booster’ cohort tends to have more men (50% vs 43%), more general Jewish people (93% vs 82%), more older people (60% vs 47% aged 70+ years), and people who were vaccinated earlier (79% vs 40% vaccinated in January). These significant differences are adjusted for when estimating protection.

**Table 1:**
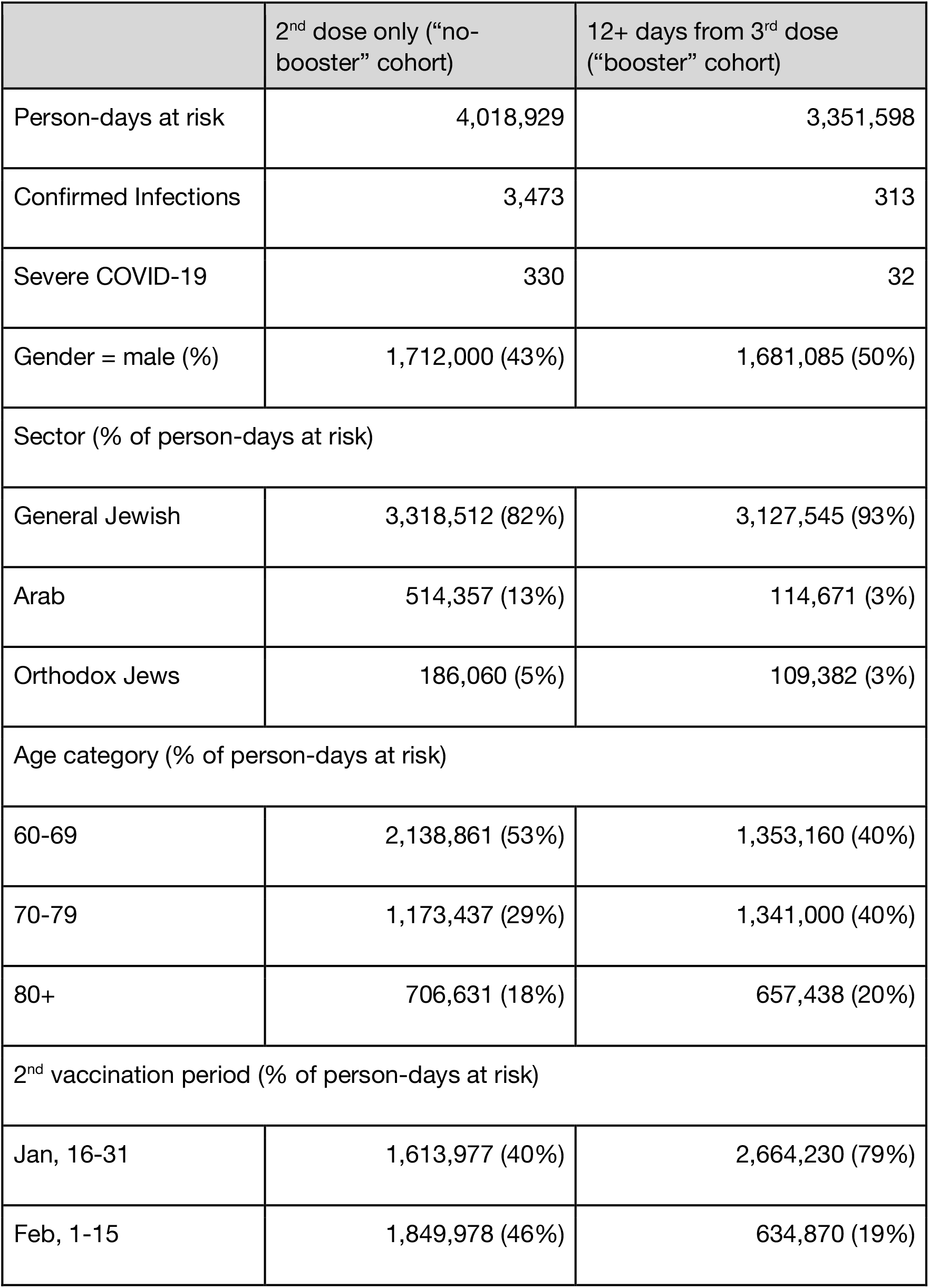
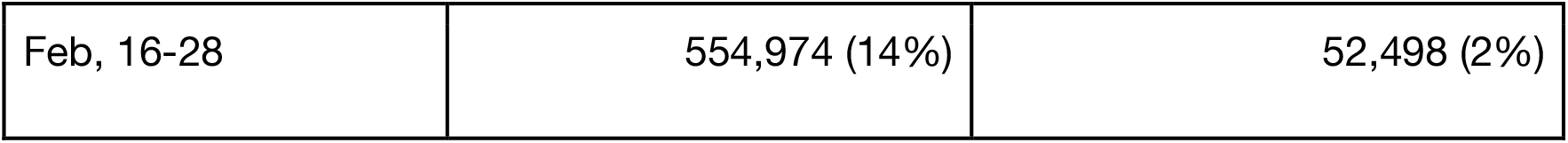
Demographic and clinical characteristics of the study population for the two study cohorts. Since the cohorts are dynamic and individuals can contribute to both cohorts, the table presents the number of person-days at risk instead of the number of individuals.

The full Poisson regression analysis for confirmed infection is given in Table S1 of the Supplementary Appendix, and the results for the booster dose protection are summarized in Table 2. The booster dose provides significant protection *–* an estimate of 11.4-fold (95% CI: [10, 12.9]) decrease in the relative risk of a confirmed infection. In the Supplementary Appendix, we provide the results of alternative analyses using matching techniques. The first analysis, following Dagan et. al.^15^, resulted in a higher estimate of the decrease in relative risk of 13.4 (95% CI: [8.2-21.4]) and the second, relying on matching by days, gave a slightly smaller estimate for the decrease in the relative risk of 9.6 (95% CI: [8.1, 11.4]).

**Table 2.**
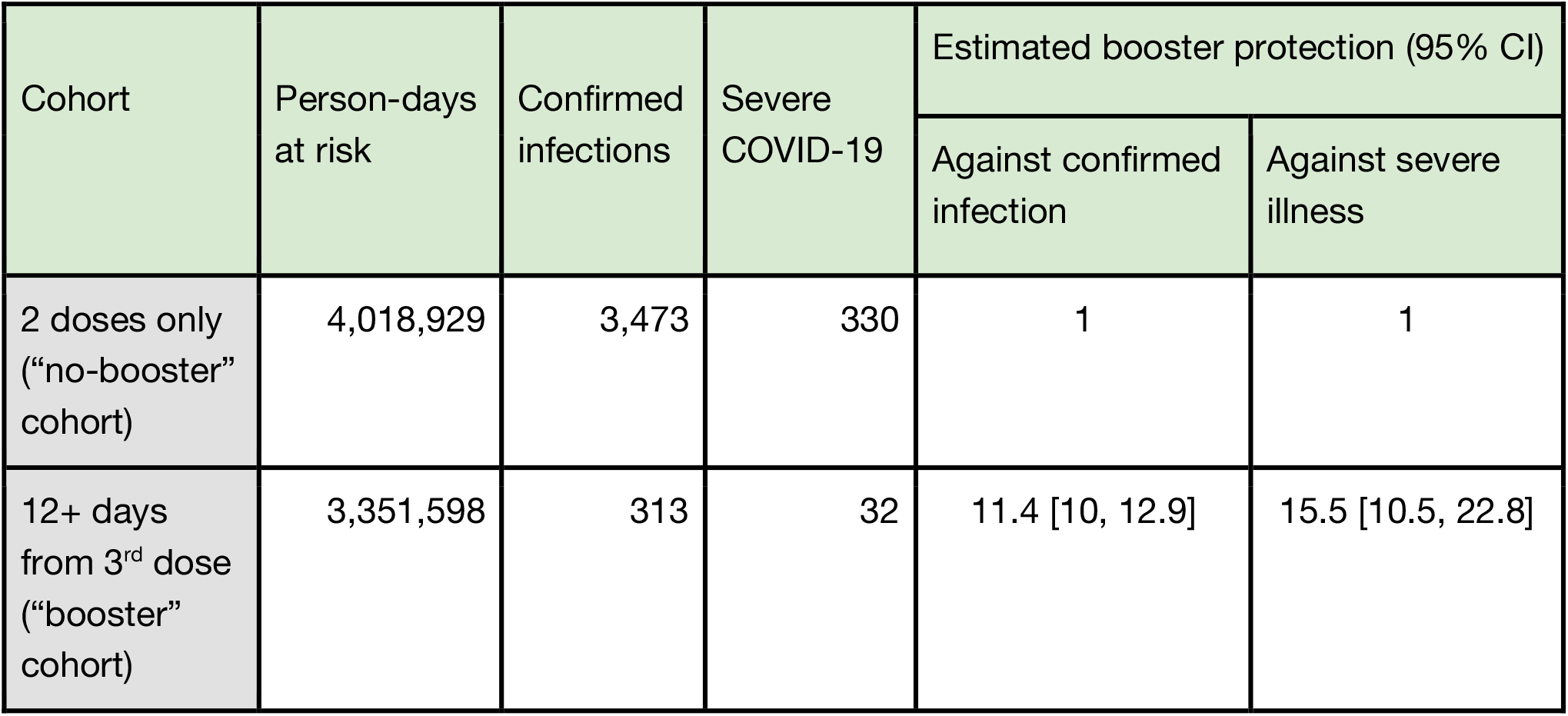
Summary of the results of the Poisson regression analysis for different cohorts: people who received only two vaccine doses and people for whom 12 days or more have passed since their booster dose. For each group, we provide the total number of person-days at risk for each cohort, the number of confirmed infections and severe COVID-19 in each cohort, and the estimated protection of the booster against confirmed infection and severe illness, given as a fold change in relative risk.

| Cohort | Person-days at risk | Confirmed infections | Severe COVID-19 | Estimated booster protection (95% CI) |  |
| --- | --- | --- | --- | --- | --- |
|  |  |  |  | Against confirmed infection | Against severe illness |
| 2 doses only ("no-booster" cohort) | 4,018,929 | 3,473 | 330 | 1 | 1 |
| 12+ days from 3<sup>rd</sup> dose ("booster" cohort) | 3,351,598 | 313 | 32 | 11.4 [10, 12.9] | 15.5 [10.5, 22.8] |

The sensitivity analysis that compared the risk of infections during 12+ days after booster to 4-6 days after booster, gave a decrease in relative risk of 4.7 (95% CI: [4, 5.4]), about half of that in the main analysis.

The protection conferred by the booster dose against severe disease also appeared high. For people with a more-than-12-days lag between the booster vaccination and severe illness, we found that the booster dose decreases the relative risk of severe disease by 15.5-fold (95% CI: [10.5, 22.8]). We validated our results using the matching-by-day analysis that gave 9.5-fold protection against severe illness (95% CI [5-19.6]).

Figure 2 presents the results of the Poisson regression analysis with the number of days after booster vaccination as additional covariates, demonstrating the protection as a function of time from the booster vaccination. After about 12 days, protection starts stabilizing at about 10-12 fold reduction in risk in line with the results presented above. As shown in Figure 2, there is an apparent protection for the booster-vaccinated group in the first days following vaccination.

**Figure 2.**
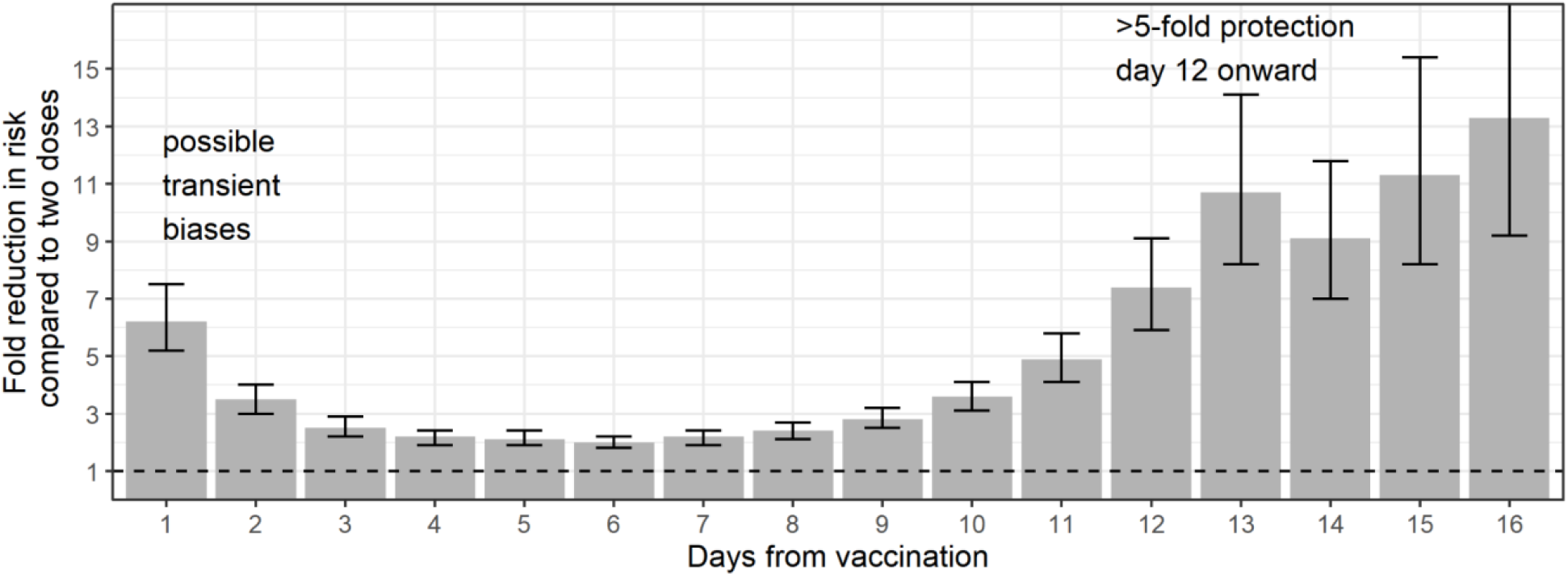
Booster protection against confirmed infection as a function of the number of days following the booster dose. Because of wide confidence intervals, only days 1-16 are shown. Protection is given as a fold reduction in risk relative to people who received only two vaccine doses. Data is based on about 1 million individuals aged 60 or older, who received the 3rd dose boost. The dashed line represents no added protection by the booster dose.

This apparent protection (i.e., days 1-4) is likely the result of the aforementioned behavioral changes that follow vaccination. As the time since vaccination progresses, the magnitude of this apparent protection decreases, indicating that the effect of behavioral change decreases. Levels of protection appear to start increasing again from day 7 post vaccination. As shown in Figure S4 these results are robust across different study periods.

## Discussion

Our analysis shows that the booster dose of the BNT162b2 vaccine is highly effective in reducing the risk of both confirmed infection and severe illness. For example, if the combined effect of waning immunity and the Delta variant decrease the efficacy of a vaccine given >6 months ago against infection to ≈50%, as recent reports have suggested ^16,17^, and if the booster dose reduces the relative risk by 10-fold, it means that the probability of a booster-vaccinated individual to being susceptible to infection would decrease to ≈5% (=50%/10) relative to unvaccinated individuals. This brings vaccine efficacy for booster-vaccinated individuals to ≈95%, similar to the original “fresh” vaccine efficacy reported against the Alpha strain ^11,15^.

On average, severe illness develops ≈5 days after the first positive-sample date (Figure S5 in the Supplementary Appendix). Thus, the follow-up period in our data is short and confidence intervals for protection against severe disease are wide. Moreover, some individuals from the booster cohort for severe illness were likely infected prior to or immediately after receiving the booster, and this could lead to an underestimate in the inferred protection against severe illness.

While our analysis attempts to address possible biases in the source data, such as the effects of confounders and behavioral changes following vaccination, there are some sources of bias that we may not correct adequately. These include differences between those receiving and not receiving the booster in care-seeking behaviors and cautiousness, and in comorbidities. Some of these possible biases are transient and fade with time since the booster vaccination, as schematically shown in Figure S1 in the Supplementary Appendix, implying that the real effectiveness of vaccination can be estimated when comparing infection rates before receiving the booster dose and after enough time has elapsed (e.g. after 12 days, Figure S1 in the Supplementary Appendix). While independent research is required in order to fully understand this behavioral model, several indications suggest that our 12 days cutoff is reasonable. First, as shown in Figure S2 in the Supplementary Appendix, people tend to perform fewer PCR tests a few days before and after the vaccination day, which is a clear source for detection bias. Over time, the number of tests that vaccinated individuals perform increases, which reduces this bias. Consistent with such behavioral change is the pattern in Figure 2, which shows a large reduction in infection risk on the first day after vaccination that monotonically decreases during the first few days, before starting to increase as the booster dose becomes effective.

Yet, confounding biases may still explain part of the observed effectiveness, and these may not disappear over time. We can put a crude lower bound on the booster efficacy by looking at time points at which the booster efficacy is not expected to be significant and behavioral differences are smaller, e.g. days 4-6 after the booster dose, and attributing the whole observed effectiveness to confounding bias. Our sensitivity analysis compared the cohort of booster-vaccinated individuals 12+ days after receiving the booster dose to the same group at days 4-6, when the booster effect is expected to be only minimally translated into a reduction in confirmed infections (Figure 2). This analysis yielded an estimate of 4.7-fold (95% CI [4.0, 5.4]) protection against confirmed infection. Even under this conservative interpretation, the demonstrated protection highlights the important role that a booster dose could play in mitigating the effects of waning immunity and immune evasion, and in mitigating the spread of VOC such as the Delta variant.

There are significant behavioral differences between the main demographic groups in Israel. Table 1 shows that the ‘booster’ cohort is highly biased toward the general Jewish population; 93% vs 82% in the ‘no-booster’ cohort. This may be a source of a selection bias that is not fully accounted for in the main analysis. We, therefore, repeated the analysis for the subset of people from the general Jewish sector. The results of the Poisson analysis were very similar to those shown in Table 2, with protection against confirmed infection of 10.9 (95% CI [9.6, 12.4]). This validates the ≈10-fold protection against infection findings of the main analysis.

Understanding the protection gained by a booster dose is critical for policy making. On July 30, 2021, Israel was the first country in the world to make available a third dose of Pfizer BNT162b2 vaccine against COVID-19 to all people aged 60 or over who had been vaccinated at least five months previously. The results of such a policy are of importance for countries that seek strategies to mitigate the pandemic. Our findings give clear indications of the effectiveness of a booster dose even against the currently dominant Delta variant.

## Data Availability

Aggregated data are given in the supplementary information. Personal data cannot be shared due to privacy.

## Ethics statement

The study was approved by the Institutional Review Board of the Sheba Medical Center. Helsinki approval number: SMC-8228-21.

## Data sharing

## Supplementary Appendix

### Supplementary Methods 1 - matching approaches

In order to validate our findings, we conducted two independent secondary analyses which rely on matching fully vaccinated individuals who received a booster dose with similar individuals who received only two vaccine doses.

The first matching approach was similar to that conducted by Dagan et.al.^15^. Briefly, each individual in the ‘booster’ cohort was matched to an individual who was in the ‘no-booster’ cohort on the booster-vaccination day based on the following characteristics: age group - (60-69,70-79 and 80+), gender, second vaccine dose week and demographic group (General Jewish, Arab, ultra-Orthodox). Follow-up for both individuals ended at the time of infection. Both individuals in a pair were censored at the end of the study or at the time the ‘no-booster’ individual got a booster dose. We calculated the probability of being free of infection in the two cohorts as a function of time using the Kaplan-Meier estimator, and compared the survival probabilities of the two cohorts at the end of the study. For each cohort, we calculated the probability of an event occurring between day 12 following the boost and the end of the study, among individuals still at risk on day 12. We used the ratio between the probabilities of the two cohorts as an estimate for the risk ratio for our population over the study time. We generated 95% confidence intervals around this estimate using the percentile bootstrap method with 100 repetitions.

A second approach matched days rather than individuals, ensuring that days in the two cohorts are comparable. Matching was performed as follows: on each day in the study period, we identified the group of individuals for whom 12 days or more passed since their booster dose (or 10 days for the severe illness analysis), and who were not previously infected (‘booster’ cohort). We randomly matched a ‘no-booster’ individual from those who received only two vaccine doses (by that same day), was not previously infected, and had the same characteristics (age group in five year window, gender, second vaccine dose week and demographic group: General Jewish, Arab, ultra-Orthodox). In order to be able to match all individuals in the ‘booster’ cohort, we conducted matching with replacement, so the same ‘no-booster’ individual could be assigned to multiple ‘booster’ individuals.

After matching was performed, we calculated the number of events (confirmed infection or severe COVID-19) occuring on the same calendar day in each of the two groups. An individual is considered severely ill at the date of first positive sample if the individual deteriorated to the corresponding condition within the study period. The ratio between the incidence of the outcomes in the ‘booster’ and ‘no-booster’ cohorts was used to estimate the marginal protection provided by the booster dose. We used non-parametric bootstrapping, with 200 bootstrap samples, followed by random matching, and reported the median ratio as the protection estimate and the 95% confidence intervals around it. Overall, out of 603,953 individuals for whom 12 days or more passed since their booster dose, 603,953 matched pairs were found.

For the first type of matching, our analysis yielded an estimate of 13.4-fold (95% CI [8.2-21.4]) for protection against confirmed infection. Due to the very small number of severe cases following the booster a reliable estimate of the protection versus severe illness using this approach was not possible.

For the second type of matching, our analysis yielded an estimate of 9.6-fold protection against confirmed infection (95% CI [8.1-11.4]) and 9.5-fold protection against severe illness (95% CI [5-19.6]). The overall agreement between the main and secondary analyses gives further reassurance that our results are robust to the employed statistical methodology.

### Supplementary Figures

**Figure S1.**
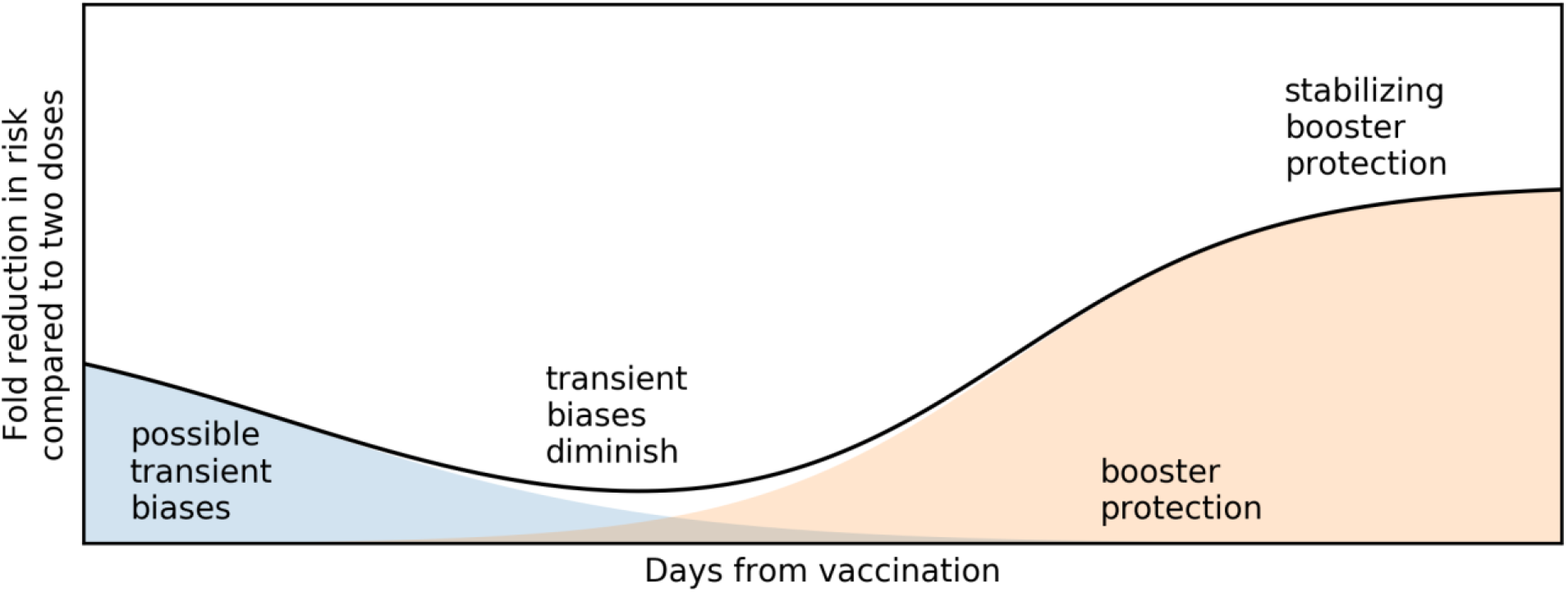
A conceptual schematic demonstrating the possible underlying dynamics of the results in Figure 2.

**Figure S2.**
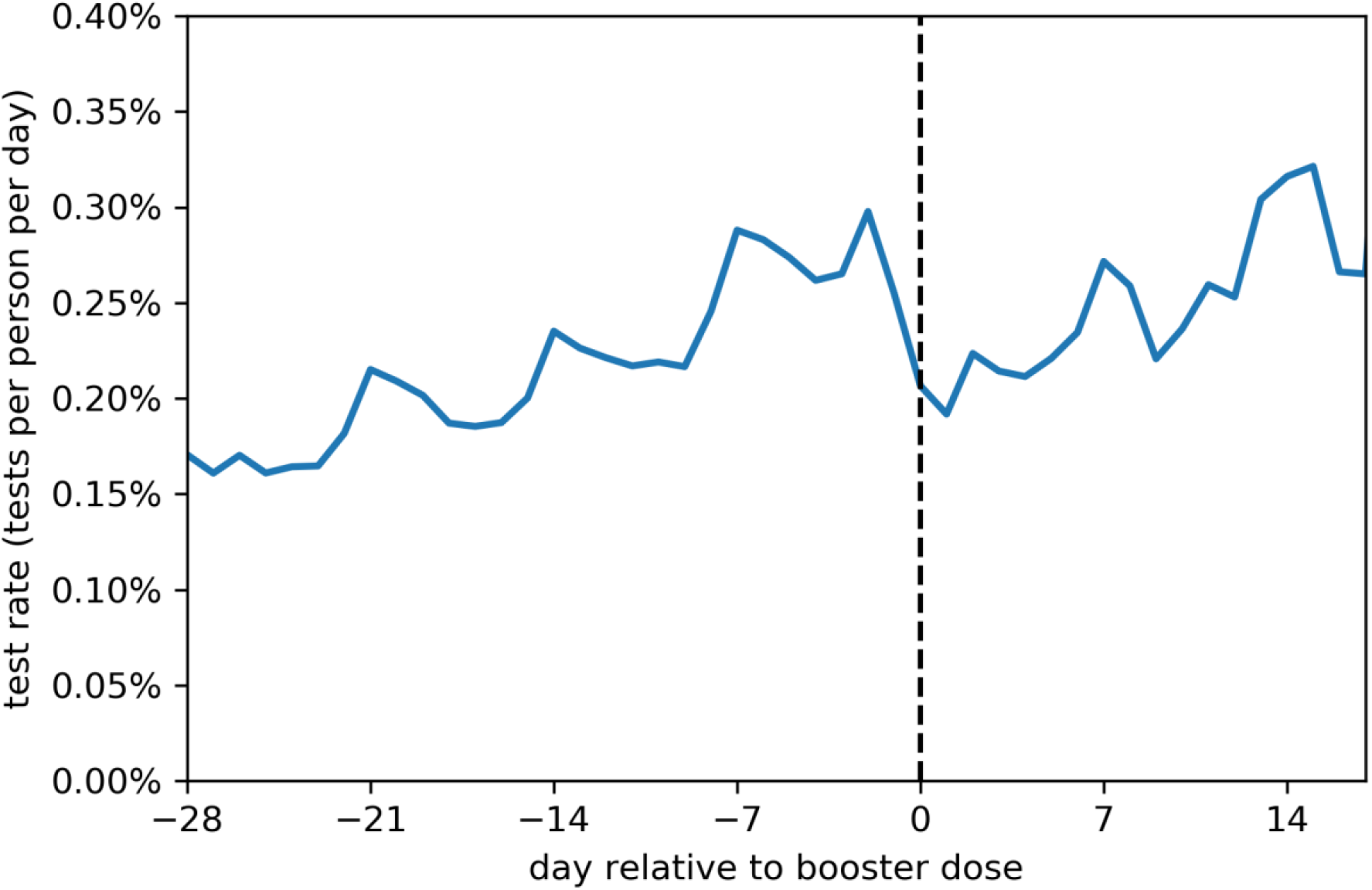
The daily rate of tests per person as a function of the time relative to the administration of the booster dose. A decrease in the rate of testing is observable just after the administration of the booster, likely reflecting transient behavioural changes in care-seeking behaviour or risk-avoidance.

**Figure S3.**
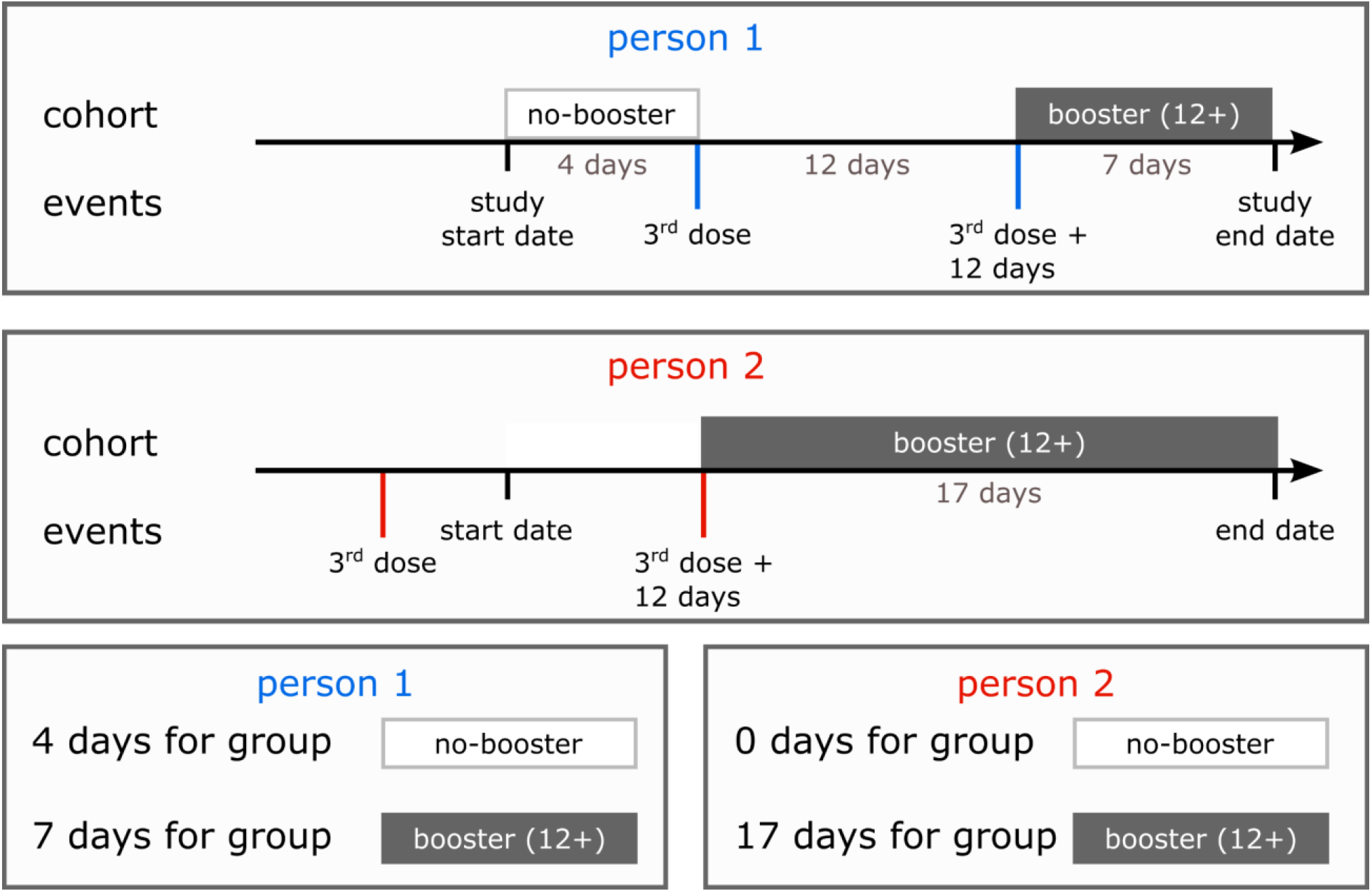
A schematic illustration of the allocation for the dynamic cohorts. We show two example timelines for two different individuals, and detail the cohort they contribute to at each point in time as well as the total days-at-risk for each person in each cohort.

**Figure S5.**
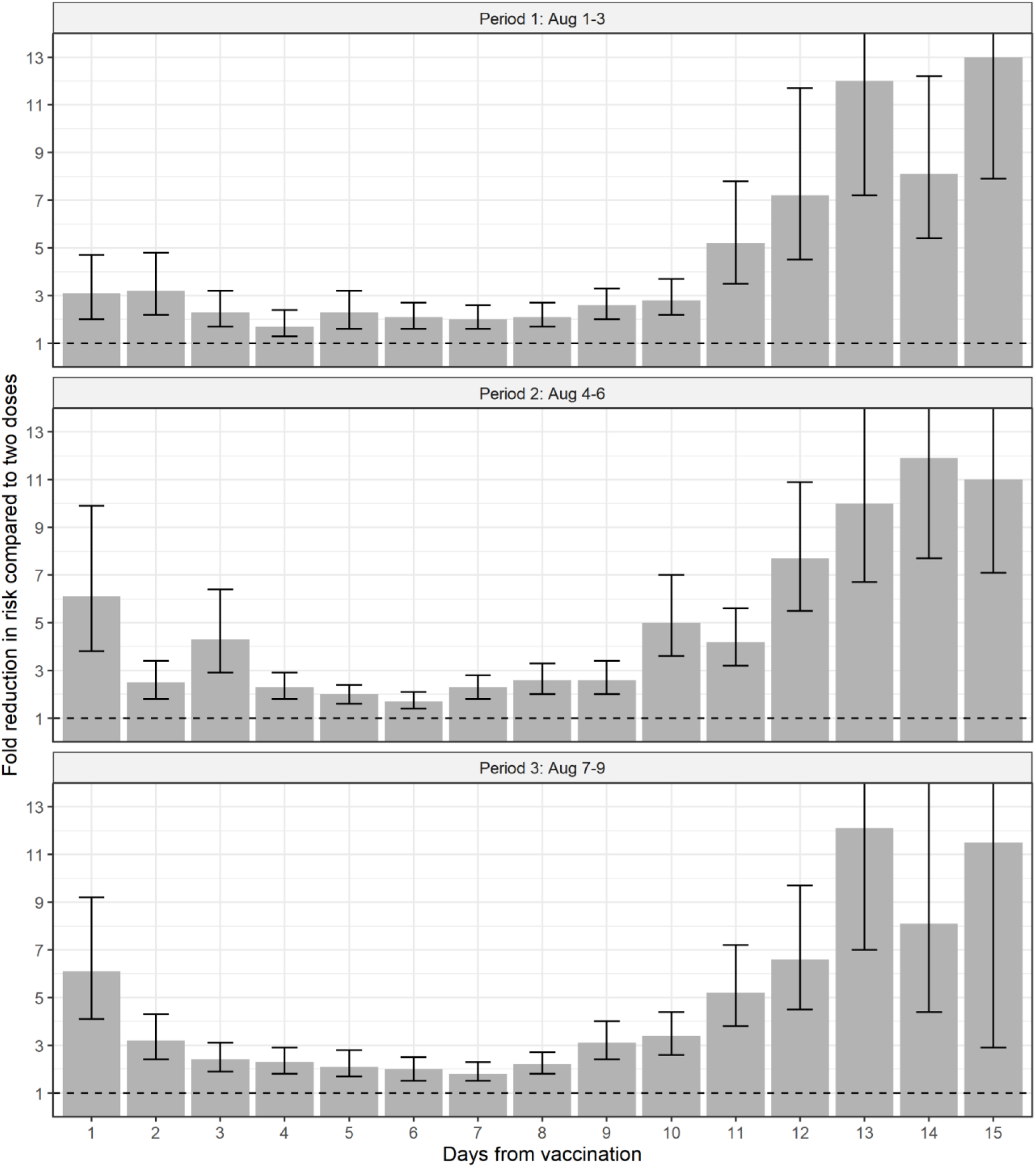
Sensitivity analysis across periods of booster vaccination for the booster protection against confirmed infection as a function of the number of days following the booster dose. Protection is given as a fold change in risk relative to people who received only two vaccine doses.

**Figure S4.**
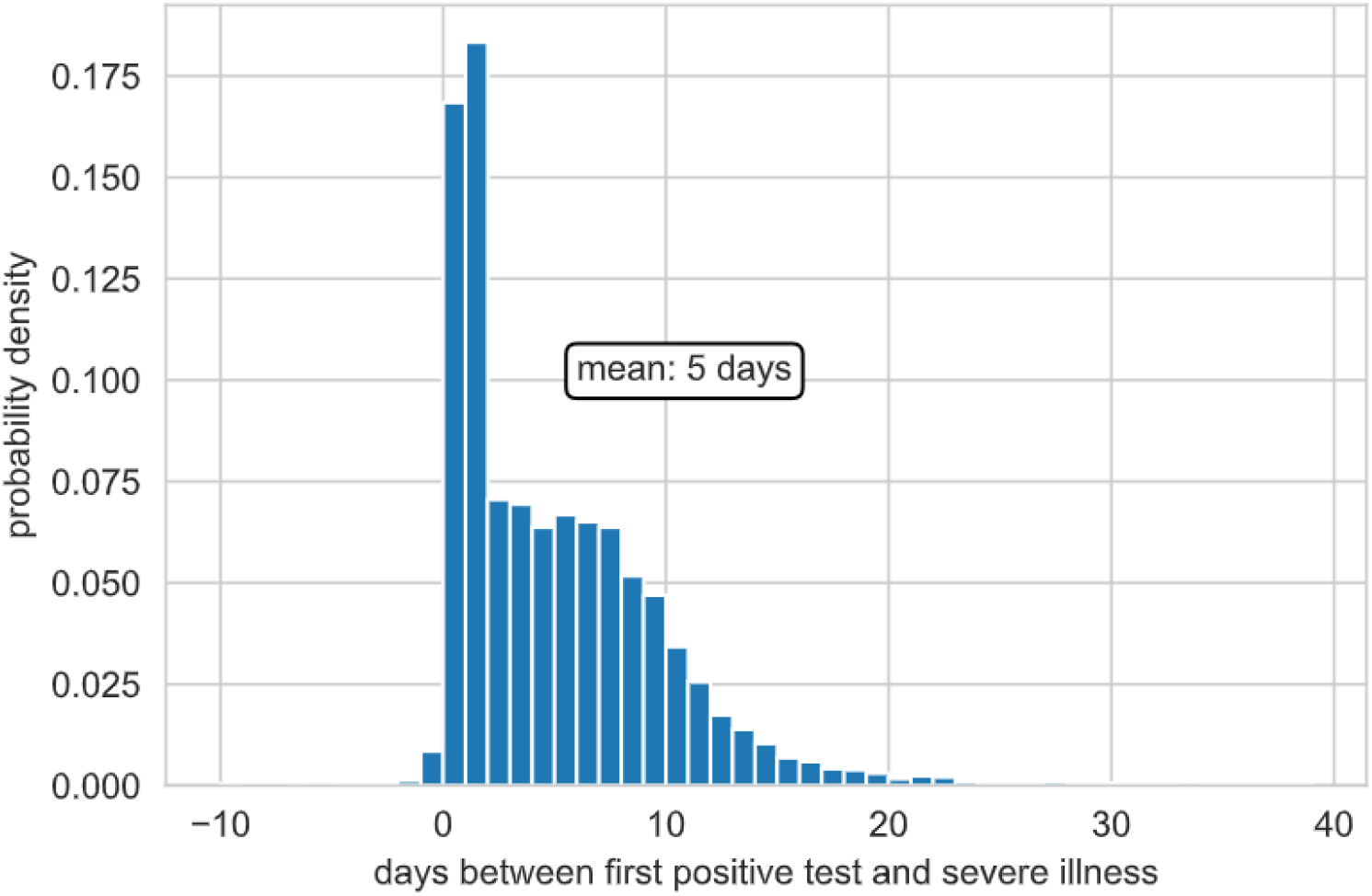
The distribution of time between first positive test and severe illness for confirmed cases between November 1st, 2020 and March 1st, 2021.

**Table S1:** Poisson regression results for confirmed SARS-CoV-2 infection.

| *term* | *estimate* | *std.error* | *statistic* | *p.value* |
| --- | --- | --- | --- | --- |
| *(Intercept)* | -6.96 | 0.06 | -124.31 | <0.001 |
| *age_category70-79* | -0.11 | 0.04 | -2.87 | 0.004 |
| *age_category80+* | -0.12 | 0.05 | -2.69 | 0.007 |
| *Gender = male* | 0.16 | 0.03 | 4.77 | <0.001 |
| *date2021-08-11* | -0.02 | 0.07 | -0.22 | 0.830 |
| *date2021-08-12* | 0.17 | 0.07 | 2.48 | 0.013 |
| *date2021-08-13* | -0.10 | 0.08 | -1.31 | 0.191 |
| *date2021-08-14* | -0.39 | 0.09 | -4.57 | <0.001 |
| *date2021-08-15* | -0.14 | 0.08 | -1.72 | 0.085 |
| *date2021-08-16* | 0.49 | 0.07 | 7.02 | <0.001 |
| *date2021-08-17* | 0.19 | 0.08 | 2.43 | 0.015 |
| *date2021-08-18* | 0.33 | 0.08 | 4.36 | <0.001 |
| *date2021-08-19* | 0.34 | 0.08 | 4.31 | <0.001 |
| *date2021-08-20* | 0.24 | 0.08 | 2.96 | 0.003 |
| *date2021-08-21* | -0.15 | 0.09 | -1.69 | 0.092 |
| *date2021-08-22* | -0.06 | 0.09 | -0.63 | 0.531 |
| *vac_periodFeb 1-15* | -0.19 | 0.04 | -5.40 | <0.001 |
| *vac_periodFeb 16-28* | -0.37 | 0.06 | -6.37 | <0.001 |
| *Sector: Arab* | -0.83 | 0.07 | -11.41 | <0.001 |
| *Sector: ultra Orthodox* | 0.30 | 0.07 | 4.51 | <0.001 |
| *Cohort 'booster'* | -2.43 | 0.06 | -38.18 | <0.001 |

**Table S2:** Poisson regression results for severe COVID-19 disease.

| *term* | *estimate* | *std.error* | *statistic* | *p.value* |
| --- | --- | --- | --- | --- |
| (Intercept) | -10.94 | 0.23 | -47.78 | <0.001 |
| age_category70-79 | 0.86 | 0.14 | 5.99 | <0.001 |
| age_category80+ | 1.79 | 0.13 | 13.26 | <0.001 |
| Gender = male | 0.96 | 0.11 | 8.67 | <0.001 |
| date2021-08-11 | 0.24 | 0.26 | 0.94 | 0.349 |
| date2021-08-12 | 0.42 | 0.25 | 1.66 | 0.097 |
| date2021-08-13 | 0.32 | 0.27 | 1.22 | 0.224 |
| date2021-08-14 | 0.11 | 0.28 | 0.40 | 0.69 |
| date2021-08-15 | 0.74 | 0.25 | 2.97 | 0.003 |
| date2021-08-16 | 0.44 | 0.27 | 1.62 | 0.106 |
| date2021-08-17 | 0.77 | 0.26 | 3.00 | 0.003 |
| date2021-08-18 | 0.48 | 0.28 | 1.69 | 0.091 |
| date2021-08-19 | 0.56 | 0.28 | 1.97 | 0.049 |
| date2021-08-20 | 0.66 | 0.28 | 2.37 | 0.018 |
| date2021-08-21 | 0.69 | 0.28 | 2.51 | 0.012 |
| date2021-08-22 | 0.95 | 0.26 | 3.62 | <0.001 |
| vac_periodFeb 1-15 | -0.31 | 0.11 | -2.78 | 0.006 |
| vac_periodFeb 16-28 | -0.84 | 0.22 | -3.77 | <0.001 |
| Sector: Arab | -0.21 | 0.19 | -1.10 | 0.273 |
| Sector: ultra Orthodox | -0.07 | 0.26 | -0.27 | 0.790 |
| Cohort 'booster' | -2.74 | 0.20 | -13.77 | <0.001 |

## References

1. Hannah Ritchie, D. B., Edouard Mathieu, Lucas Rodés-Guirao, Cameron Appel, Charlie Giattino, Esteban Ortiz-Ospina, Joe Hasell, Bobbie Macdonald & Roser, M. Coronavirus Pandemic (COVID-19). Our World Data (2020).

2. Goldberg, Y. et al. Waning immunity of the BNT162b2 vaccine: A nationwide study from Israel.

3. Investigation of SARS-CoV-2 variants of concern: technical briefings. GOV.UK https://www.gov.uk/government/publications/investigation-of-novel-sars-cov-2-variant-variant-of-concern-20201201.

4. Mizrahi, B. et al. Correlation of SARS-CoV-2 Breakthrough Infections to Time-from-vaccine; Preliminary Study. 2021.07.29.21261317 https://www.medrxiv.org/content/10.1101/2021.07.29.21261317v1 (2021) doi:10.1101/2021.07.29.21261317.

5. Wall, E. C. et al. Neutralising antibody activity against SARS-CoV-2 VOCs B.1.617.2 and B.1.351 by BNT162b2 vaccination. The Lancet 397, 2331–2333 (2021).

6. Pfizer Inc. - Pfizer Quarterly Corporate Performance – Second Quarter 2021. https://investors.pfizer.com/events-and-presentations/event-details/2021/Pfizer-Quarterly-Corporate-Performance--Second-Quarter-2021/default.aspx.

7. Khoury, D. S. et al. Neutralizing antibody levels are highly predictive of immune protection from symptomatic SARS-CoV-2 infection. Nat. Med. 27, 1205–1211 (2021).

8. Lustig, Y. et al. BNT162b2 COVID-19 vaccine and correlates of humoral immune responses and dynamics: a prospective, single-centre, longitudinal cohort study in health-care workers. Lancet Respir. Med. 0, (2021).

9. Walsh, E. E. et al. Safety and Immunogenicity of Two RNA-Based Covid-19 Vaccine Candidates. N. Engl. J. Med. 383, 2439–2450 (2020).

10. Muhsen, K. et al. A nationwide analysis of population group differences in the COVID-19 epidemic in Israel, February 2020-February 2021. Lancet Reg. Health Eur. 7, 100130 (2021).

11. Haas, E. J. et al. Impact and effectiveness of mRNA BNT162b2 vaccine against SARS- CoV-2 infections and COVID-19 cases, hospitalisations, and deaths following a nationwide vaccination campaign in Israel: an observational study using national surveillance data. The Lancet 397, 1819–1829 (2021).

12. McAloon, C. et al. Incubation period of COVID-19: a rapid systematic review and meta-analysis of observational research. BMJ Open 10, e039652 (2020).

13. Xin, H. et al. The Incubation Period Distribution of Coronavirus Disease 2019: A Systematic Review and Meta–analysis. Clin. Infect. Dis. (2021) doi:10.1093/cid/ciab501.

14. R Core Team. R: A Language and Environment for Statistical Computing. (R Foundation for Statistical Computing, 2020).

15. Dagan, N. et al. BNT162b2 mRNA Covid-19 Vaccine in a Nationwide Mass Vaccination Setting. N. Engl. J. Med. 384, 1412–1423 (2021).

16. Puranik, A. et al. Comparison of two highly-effective mRNA vaccines for COVID-19 during periods of Alpha and Delta variant prevalence. 2021.08.06.21261707 https://www.medrxiv.org/content/10.1101/2021.08.06.21261707v2 (2021) doi:10.1101/2021.08.06.21261707.

17. Tang, P. et al. BNT162b2 and mRNA-1273 COVID-19 vaccine effectiveness against the Delta (B.1.617.2) variant in Qatar. 2021.08.11.21261885 https://www.medrxiv.org/content/10.1101/2021.08.11.21261885v1 (2021) doi:10.1101/2021.08.11.21261885.

